# Causal effects of senescence through intrinsic epigenetic age acceleration on age-related diseases: a Mendelian randomization study

**DOI:** 10.1101/2020.04.27.20081406

**Authors:** F. Gatto, P. Ueda

## Abstract

Older age is associated with many diseases. However, it is unclear whether the biological mechanisms underpinning senescense (biological ageing) is a cause of age-related diseases (ARDs). In this Mendelian randomization study, we examined the causal effects of intrinsic epigenetic age acceleration (IEAA) - a process assumed downstream of senescence - on the occurrence of ARDs. In a metanalysis of 18 genome-wide association studies (GWAS) spanning 12 ARDs in ~27 million people of European ancestry (median per GWAS: 17’008 cases vs. 40’940 controls), IEAA-associated SNPs did not significantly increase the risk of ARDs as a group (odds-ratio (OR) = 0.99) nor of any individual ARD (OR range: 0.87-1.04). However, one SNP, rs2736099 on TERT, was significantly associated with 8 of 10 ARDs with increased risk for lung and ovarian cancer, atrial fibrillation, and benign prostatic hyperplasia and lower risk for Alzheimer’s disease, rheumatoid arthritis, cardiovascular artery disease and breast cancer. No associations between rs2736099 and known ARD risk factors were found strengthening the causal role of rs2736099-associated senescence. While IEAA was not found as a unifying cause of ARDs, senescence seems to cause some ARDs while preventing others through a mechanism only partially affecting IEAA and associated with rs2736099.

## Introduction

Old age is the main risk factor for many diseases, including cardiovascular diseases, neurodegenerative diseases and cancer. These diseases alone account for one third of the disease burden in high sociodemographic index countries (1). Geroscience is an emerging research area that studies the link between age-related diseases (ARDs) and senescence - defined as the intrinsic process that determines the molecular and physiological changes resulting in the progressive and cumulative deterioration of body functions and culminating in death. The underlying theory of geroscience is that ARDs can be prevented and health-span extended by targeting senescence (2). However, despite the strong association between the two, and the striking overlap between the hallmarks of biological ageing and some mechanisms at the origin of ARDs, it remains uncertain whether senescence is a unifying cause of ARDs (3,4). Importantly, if causality were to be demonstrated, the rationale to focus resources on targeting senescence to eradicate ARDs rather than focusing on each disease individually is obvious.

To assess whether senescence causes ARDs is challenging. Decades of molecular mechanistic studies to test this hypothesis were performed in model organisms (3,5,6). Clinical evidence is emerging but still limited (7). While epidemiological studies have linked senescence to multiple ARDs (8), their observational design preclude conclusions regarding causality as varying levels of senescence cannot be randomized and any observed associations with ARDs may be caused by other factors. In this context, Mendelian randomization (MR) is a study design that, under certain assumptions, can demonstrate causality (9). In brief, MR takes advantage of the fact that a population is randomly divided in two groups at conception to bear a certain genetic variant or not. As long as the genetic variant is not directly or indirectly associated with ARDs (if not via senescence), it is possible to select a variant - typically a single nucleotide polymorphism (SNP) - such that one group is more exposed to senescence than the other. If the exposed group is subsequently observed to feature higher incidence of ARDs than the unexposed group, one can infer that senescence is a cause of ARDs. Conversely, if no difference is observed, senescence is likely not a significant cause of ARDs.

Senescence is a complex biological process unfolding through many inter-related pathways, e.g. from insulin and mTOR signaling to oxidative stress and cellular senescence, collectively representing the hallmarks of aging (10). No biomarker is universally accepted as a measure of senescence. In the last decade, DNA methylation at specific CpG sites was shown to effectively parallel the genetic and environmental factors affecting biological age across the life course. Epigenetic clocks are defined as the weighted average of methylation in the clock-specific set of CpG sites. An example of these is Horvath’s clock that first reported a very strong correlation with chronological age (Pearson correlation coefficient = 0.96) and the difference between an individual’s chronological age and the predicted age was recently shown to recapitulate longevity in a number of studies (11–15). This difference is also referred to as intrinsic epigenetic age acceleration (IEAA) since it measures cell-intrinsic methylation changes that are consistent across tissues and show weak association with lifestyle factors (16). It is therefore plausible to hypothesize that a) IEAA is a process controlled solely by senescence and b) IEAA can serve as a biomarker of senescence.

In this study, we tested causality between senescence and 12 ARDs - here broadly defined as diseases associated with older age - using two-sample MR with IEAA as exposure and a set of single nucleotide polymorphisms (SNPs) recently associated with IEAA as instrumental variables (17) under the assumption that IEAA is a process downstream of senescence. We enumerated three different scenarios modeled through directed acyclic graphs and tested using MR (Figure 1): 1) IEAA-associated SNPs are not associated with ARDs (individually or as a group), i.e. senescence does not cause ARDs; 2) IEAA-associated SNPs are associated with ARDs, i.e. senescence causes ARDs; 3) one or more IEAA-associated SNPs (yet not all) are associated with ARDs, i.e. senescence causes ARDs but through a mechanism only partially affecting IEAA. A fourth scenario is that senescence causes ARDs but does not control IEAA, which would violate our assumption.

**Figure 1.**
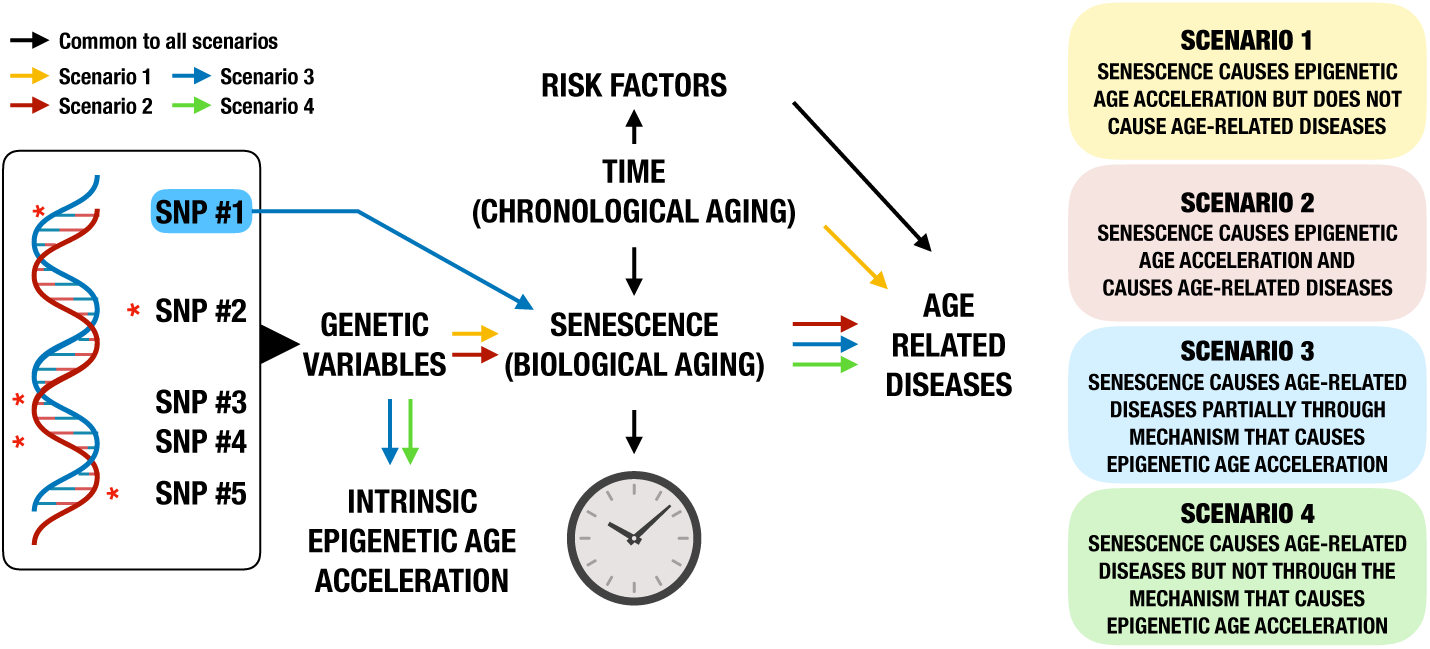
Directed acyclic graphs used in the present Mendelian randomization (MR) study to test different scenarios for causality between senescence - assumed to control IEAA (except in scenario 4) - and ARDs.

## Results

### Study design and population

As instrumental variables for two-sample MR, we used 9 SNPs previously correlated with IEAA: rs388649, rs6440667, rs79070372, rs1011267, rs62078811, rs4712953, rs10778517, rs76244256, and rs79070372 (17). Next, we used MR-Base (18) - a curated database of complete GWAS results - to assemble 18 independent GWAS on 12 distinct ARDs (Alzheimer’s disease, atrial fibrillation, benign prostatic hyperplasia, breast cancer, coronary heart/artery disease, ischemic stroke, lung cancer, osteoarthritis, ovarian cancer, pancreatic cancer, Parkinson’s disease and rheumatoid arthritis) from populations of European ancestry (Table S1). On median, the sample size was 57 284 subjects (range: 1 489 - 1 030 836) with 17 008 cases (range: 753 - 76 192) and 40 941 controls (range: 736 - 970 216). Of the 9 IEAA-associated SNPs, a median 7 SNPs were used as instrumental variables in each GWAS (range: 1 - 8), of which a median 0 SNPs (range: 0–3) were replaced with linkage disequilibrium (LD) proxies (Table S1). (During harmonization, one or more SNPs were omitted due to ambiguous minor allele frequency for palindromic alleles, e.g. rs388649 was omitted in all GWAS.)

### Mendelian randomization using IEAA-associated SNPs

We performed Mendelian randomization using RAPS (Robust Adjusted Profile Score, (19)) - a method devised to combine multiple weak instruments and to correct for systematic and idiosyncratic pleiotropy observed in real datasets violating MR assumptions. In a random effects metanalysis model of 17 GWAS, IEAA-associated SNPs were not significantly correlated with ARDs as a group (OR = 0.995, 95% CI [0.987 - 1.003], p = 0.21, Table S2, Figure S1 - note that 1 GWAS for benign prostatic hyperplasia was omitted due to its excessive weight in the metanalysis with no remarkable loss of information). In 16 of 18 GWAS, encompassing 11 of 12 ARDs, we did not observe any statistically significant association between IEAA-associated SNPs and the individual ARD (Figure 2). In 2 of 3 GWAS on rheumatoid arthritis, we observed a significant protective effect of IEAA (OR range: 0.87-0.96 Table S2).

**Figure 2.**
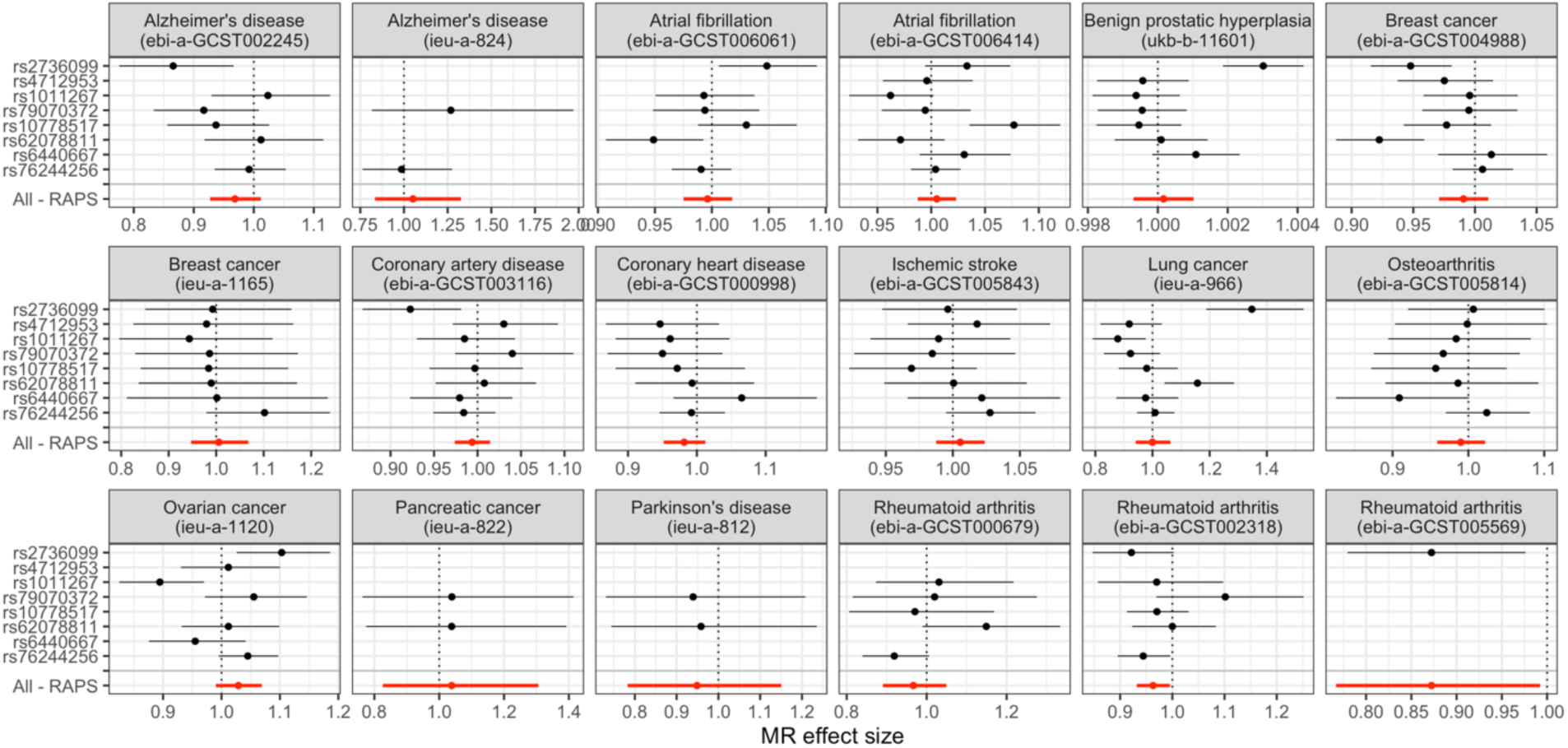
Forest plot of up to 8 IEAA-associated SNPs in 18 GWAS spanning 12 ARDs. MR effect sizes in terms of OR is shown as a point for each individual SNPs and all combined as weak instruments using RAPS (in red) with 95% CI as error bar

### Mendelian randomization using rs2736099

We detected elevated heterogeneity for IEAA-associated SNPs in 6 of 18 GWAS from the analysis above (Q-statistics = 12.9 - 39.2, *p <* 0.05). This seemed to be driven by one SNP, rs2736099 (Figure 2). We therefore performed a two-sample Mendelian randomization analysis using Wald ratio on rs2736099 in 10 distinct ARDs across 13 GWAS. Whereas a metanalysis of all exposure-outcome pairs revealed no overall significant association between rs2736099 and ARDs as a group (OR = 0.997, 95% CI [0.940 - 1.057], p > 0.05), we detected statistically significant associations in 8 of 10 individual ARDs (Table 1). We sub-grouped ARDs into “senescence-protected”, “senescence-induced” and “senescence-independent” groups based on a *post-hoc* analysis of the MR effect sizes and performed a metanalysis per sub-group (Figure 3). We recovered a causal effect in 4 ARDs in the “senescence-induced” group, comprising atrial fibrillation, lung and ovarian cancer, and benign prostatic hyperplasia (OR = 1.088, 95% CI [0.988 - 1.198], p = 0.086 with elevated heterogeneity (I^2^ = 88%)), a significant protective effect for 4 ARDs in the “senescence-protected” group, comprising Alzheimer’s disease, breast cancer, coronary artery disease (CAD) and rheumatoid arthritis (OR = 0.932, 95% CI [0.908 - 0.958],, p < 10^−6^) and no effect in the “senescence-independent” groups, comprising ischemic stroke or osteoarthritis. Sub-group differences were statistically significant (Q = 14.1, p = 0.009).

**Table 1.** Odds-ratio (OR) estimates and 95% confidence interval (CI) for each exposure-outcome pair (e.g. rs2736099 vs. individual ARD) in a two-sample MR tests using Wald ratio.

| ARD | Subgroup | OR | 95% CI | p |
| --- | --- | --- | --- | --- |
| Alzheimer's disease (ebi-a-GCST002245) | Senescence-protected | 0.866 | (0.776-0.966) | 0.010 |
| Rheumatoid arthritis (ebi-a-GCST002318) | Senescence-protected | 0.922 | (0.847-1.003) | 0.057 |
| Coronary artery disease (ebi-a-GCST003116) | Senescence-protected | 0.923 | (0.868-0.981) | 0.010 |
| Breast cancer (ebi-a-GCST004988) | Senescence-protected | 0.948 | (0.915-0.981) | 0.002 |
| Rheumatoid arthritis (ebi-a-GCST005569) | Senescence-protected | 0.872 | (0.779-0.976) | 0.017 |
| Breast cancer (ieu-a-1165) | Senescence-protected | 0.993 | (0.851-1.158) | 0.926 |
| Atrial fibrillation (ebi-a-GCST006061) | Senescence-induced | 1.048 | (1.006-1.092) | 0.024 |
| Atrial fibrillation (ebi-a-GCST006414) | Senescence-induced | 1.033 | (0.994-1.074) | 0.095 |
| Ovarian cancer (ieu-a-1120) | Senescence-induced | 1.103 | (1.026-1.186) | 0.008 |
| Lung cancer (ieu-a-966) | Senescence-induced | 1.348 | (1.188-1.529) | 3.60E-06 |
| Benign prostatic hyperplasia (ukb-b-11601) | Senescence-induced | 1.003 | (1.002-1.004) | 2.95E-07 |
| Osteoarthritis (ebi-a-GCST005814) | Senescence-independent | 1.007 | (0.921-1.101) | 0.886 |
| Ischemic stroke (ebi-a-GCST005843) | Senescence-independent | 0.996 | (0.947-1.048) | 0.884 |

**Figure 3.**
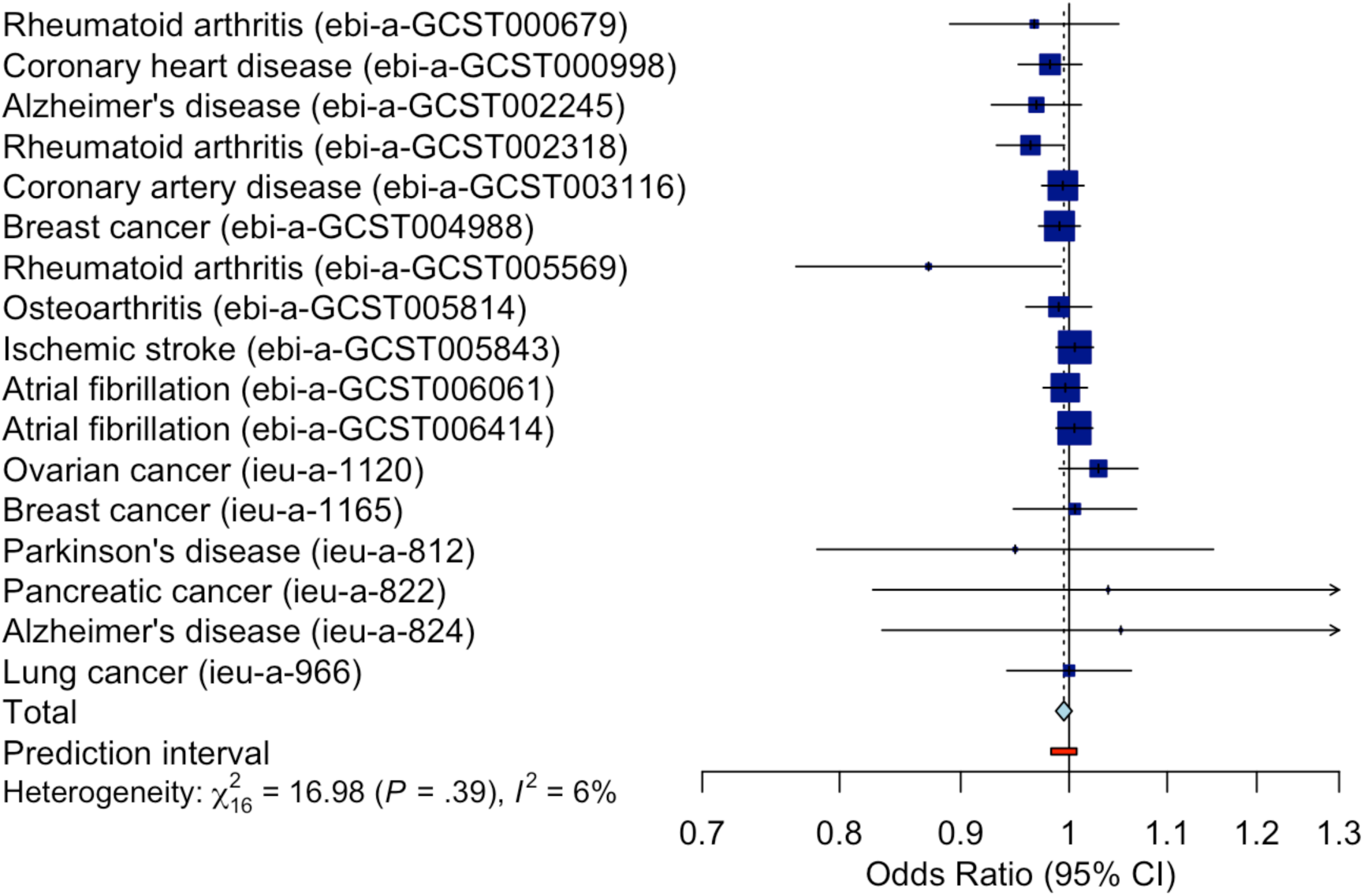
Forest plot of rs2736099-associated MR effect sizes (in terms of ORs) for 13 GWAS spanning 10 ARDs (one point for each GWAS) and when metanalysed per sub-group and overall using a random effect model. The bars represent the corresponding 95% CI.

We checked whether rs2736099 predisposed to 14 hereditary, lifestyle-related, physiological and physical risk factors for ARDs (N = 39 GWAS) that may in principle either cause ARDs bypassing senescence or cause IEAA without requiring senescence, thereby violating the study assumptions (Table S3, Figure S2–3). rs2736099 correlated with lower LDL cholesterol (OR = 0.98, 95% CI [0.96 - 1.00], p = 0.02, Figure S3), concordant with the above-mentioned protective effect on CAD. Otherwise, rs2736099 was not significantly associated with any of the other 13 risk factors. In addition, we did not recover a genome-wide association between rs2736099 and any traits other than IEAA in the NHGRI-EBI GWAS Catalog, which includes 601 reportedly unique traits (20).

## Discussion

The role of biological ageing, or senescence, in the development of ARDs has attracted significant debate for its important societal implications (2,21). Certain interventions, for example targeting the tissue accumulation of senescent cells - a mechanism directly controlled by biological ageing, successfully prolonged mice health-span (22). From the present study, we conclude that senescence is causally implicated in most ARDs through a mechanism partially captured by IEAA (scenario 3 from Figure 1) based on the following observations: rs2736099 correlates with IEAA and with most ARDs but not with known risk factors; and rs2736099 can be reasonably assumed to affect ARDs only through IEAA, which is itself exclusively controlled by senescence. If rs2736099 is truly randomly assigned in the study population, then it descends that senescence should cause most ARDs. At the same time, this study does not support the notion that IEAA is a) either a unifying cause of ARDs or b) exhaustively represents senescence (otherwise all 8 SNPs associated with IEAA - and not only rs2736099 - would have correlated with ARDs, scenario 2). Paradoxically, senescence seemed to have a protective effect in some ARDs, a case made stronger for CAD wherein LDL cholesterol - a causal risk factor of CAD - was also decreased in association with rs2736099.

Here, rs2736099 was used as instrumental variable for MR. However, being the only IEAA-associated SNP through which senescence was causally implicated in ARDs, we hypothesize that it may participate in ageing mechanisms causing ARDs. This is also motivated by the fact that rs2736099 resides in TERT, a key gene for cellular immortality (23). rs2736099 lies in an intronic region not extensively investigated. Lu et al. (24) first described the association of rs2736099 with IEAA and experimentally showed that hTERT induction in primary fibroblasts prevented senescence in vitro. In a study by Bojesen et al. (25), various SNPs in TERT independently associated with risk of ARDs (specifically neoplasms). Consistent with our results, these SNPs correlated with increased risk of ovarian cancer and decreased risk of breast cancer. The present study raises more questions on the role of TERT SNPs in senescence. Lu et al. (24) themselves noted rs2736099’s surprising effect on IEAA considering that telomere maintenance by TERT supposedly decelerate - and not accelerate, as observed - epigenetic age.

Limitations in this MR study include the potential violation of its stringent assumptions. Primarily, rs2736099 may be associated with ARDs through unknown factors, bypassing senescence, thereby questioning the causal link. The only illuminating experiment to refute this possibility is described by Lu et al., in which hTERT induction resulted in IEAA, strongly implicating IEAA in rs2736099 mechanism of action (24). Secondly, the inability to show that IEAA causes ARDs (scenario 1) could be due to low statistical power as illustrated by the case of benign prostatic hyperplasia with its minute but highly significant association with rs2736099; or that some of the previously reported associations between SNPs used in our study and IEAA were based on spurious findings; or that the most representative IEAA-associated SNPs are yet to be discovered. Thirdly, while all outcomes selected for this study were chosen because they were associated with older age, there is no clear consensus of how ARDs should be defined (26). However, the study conclusions are unlikely to change by including or excluding some diseases, as the diseases included in this study comprise some of the most common ARDs.

In conclusion, these findings support that a) mechanisms of senescence that control IEAA are not a unifying cause of ARDs; yet b) interventions aimed at eliminating senescence are likely to impact the occurrence of ARDs, with the paradoxical addendum that c) these may lead to the prevention of some ARDs while inducing others. Further understanding of the biological mechanisms behind senescence may identify those controlling senescence-induced ARDs.

## Methods

### Data sources and eligibility

For the exposure and its instrumental variables, the 10 SNPs associated with IEAA in populations of European ancestry in Gibson et al. 2019 (17) were used (Table S1). Of these, one SNP, rs7744541, was omitted due to high linkage disequilibrium (LD).

For the outcomes, MR-Base (https://gwas.mrcieu.ac.uk/) was last accessed in March 2020 (18). The following (and immediately related) terms for ARDs were searched as traits: “Alzheimer’s disease”, “arthritis”, “atrial fibrillation”, “cardiovascular disease”, “cancer”, “cataract”, “coronary artery/heart disease”, “COPD”, “dementia”, “heart failure”, “hyperplasia”, “macular degeneration”, “myocardial infarction”, “neoplasm”, “osteoporosis”, “Parkinson’s disease”, “stroke”. The search yielded 43 GWAS (Table S5). Of these, 17 had non independent populations or suspected so. Of the remaining 26, 7 with mixed or non European ancestry and one with less than 750 cases were excluded, yielding 18 GWAS included in the study.

### Mendelian randomization analysis

The 9 SNPs were extracted from each individual GWAS. If the SNP was not genotyped in a certain GWAS, LD proxies (R^2^ > 0.80) were used. If palindromic, the minor allele frequency (MAF) was capped to 0.30. If no LD proxies were found, or if palindromic and MAF > 0.30, then the SNP was omitted in that GWAS to avoid potentially wrong inferences (27). In 10 of 18 GWAS, all 9 SNPs were found; in four of 18 GWAS, 8 of 9 SNPs were found; while in the remaining four GWAS, four or less SNPs were found. LD proxies were used in 8 of 18 GWAS, with rs388649 and rs79070372 being the most commonly replaced SNPs (in 100% and 75% of these 8 GWAS, respectively). The outcome and exposure datasets were harmonized to be relative to the same effect allele. We inferred positive strand alleles using allele frequencies for palindromes. One or more SNPs were removed because palindromic with ambiguous allele frequency, e.g. rs388649 was discarded in all GWAS. Harmonized data is presented in Table S6. The mean F-statistics in the regression of the exposure to the instrumental variables was equal to 44 if all 8 IEAA-SNPs were used (in the other cases, the F statistics ranged between 34 - 54), indicating low weak instrument bias (28,29).

We performed two-sample MR test using RAPS (Robust Adjusted Profile Score, (19)) for each exposure-outcome pair (IEAA vs. individual ARD). The effect size and standard error for each exposure-outcome pair was used in a random effects model to estimate the meta-effect size for all ARDs as a group using the inverse variance method, the Empirical Bayes estimator for τ^2^, and the Q-profile method for the confidence interval of τ^2^. As a measure of between-study heterogeneity, we computed a *I*^2^ = 6%, indicating low heterogeneity. The weight of each source ranged from 0.1% to 16.3% after excluding ukb-b-11601, which would otherwise carry 96.7% weight given the substantially larger sample size. Excluding ukb-b-11601 did not remarkably affect meta-effect size estimates.

We performed a number of sensitivity analyses. Heterogeneity Q-statistics was obtained for each exposure-outcome pair using the inverse-variance wight method. In 12 of 18 GWAS, no statistically significant heterogeneity was observed (p > 0.05). In 6 of 18 GWAS, in particular in benign prostatic hyperplasia (ukb-b-11601) and lung cancer (ieu-a-966), and to a lesser extent in atrial fibrillation (ebi-a-GCST006061 and ebi-a-GCST006414), in breast cancer (ebi-a-GCST004988) and in ovarian cancer (ieu-a-1120), we observed statistically significant heterogeneity (Q statistics range: 12.9 - 39.2). After inspection of the results (Figure 2), we deemed these effects not to reject the conclusions and to motivate the choice to focus on rs2736099. Horizontal pleiotropy was tested using Egger regression for each exposure-outcome pair (except in 4 GWAS where 4 or less SNPs were used). We observed no statistically significant intercept estimates (p > 0.05 for all), although the use of few instrumental variables suggests that the test is likely under-powered. Nevertheless, we observed that the nominal intercept estimates were small (range: −0.05 to 0.06). Finally, we used the Steiger test to check the validity of the assumption that exposure causes outcome. In all cases, we observed a statistically significant correct causal direction (p < 10^−11^ for all exposure-outcome pairs).

We performed MR as described above using rs2736099 as the only instrumental variable. The SNP was genotyped in 13 GWAS (no LD proxies were used) and instead of RAPS, the Wald ratio method was used. Sub-groups for the meta-analysis were derived by categorizing ARDs based on a post-hoc analysis of the MR effect size: ARDs with low non-significant effect size (|beta| < 7·10^−3^, p > 0.05) as “senescence-indepedent”, ARDs with high or significant positive effect size (beta > 7·10^−3^) as “senescence-induced”, and ARDs with high or significant negative effect size (beta < −7·10^−3^) as “senescence-protected”. The meta-analysis was performed as described above for each sub-group separately as well as collectively. Sub-group differences were tested using a random effects model.

We searched for risk factors from traits in MR-Base (last accessed March 2020). A total of 39 GWAS from populations of European ancestry encompassing hereditary factors (illnesses of mother/father), lifestyle-related factors (sleep, physical activity, social connections, alcohol, air pollution, socioeconomic status, smoking, and stress), physiological and physical factors (diabetes, obesity, hyperlipidaemia, inflammation, and hypertension) were retrieved (at least 2 GWAS per risk factor). We extracted the regression coefficient beta and the standard error in association with rs2736099 and metanalysed all GWAS corresponding to one risk factor (for a total of 14 risk factors) using a random effects model (Table S3). Other genome-wide associations with rs2736099 were searched among traits in the NHGRI-EBI GWAS Catalog (last accessed March 2020) (19).

Odds-ratio estimates and 95% confidence interval were computed by exponentiating the regression coefficient beta and its standard error. Note that these estimates are (more or less) biased for non-linear models (30) but should still uphold the conclusion on causality of the effects for at least part of the study population regardless of their magnitude.

All analyses were performed in R (version 3.6.3) using the R-packages *TwoSampleMR* (for two-sample Mendelian randomization, (18)) and *meta* (for GWAS metanalysis, (31)) with default parameters except where stated otherwise.

### Code availability

Analyses in this study can be accessed at https://github.com/gattofr/SenescenceMR

## Data Availability

Analyses in this study can be accessed at https://github.com/gattofr/SenescenceMR

## Acknowledgements

The authors wish to acknowledge Raphael Ferreira (Harvard Medical School, Boston, MA) for critical reading and help with the figures, Riccardo Burioni (University of Edinburgh, Edinburgh, UK) and Gibran Hemani (University of Bristol, Bristol, UK) for useful suggestions. The authors declare no conflicts of interest. The study received no specific funding.

## Supplementary Material

**Figure S1.**
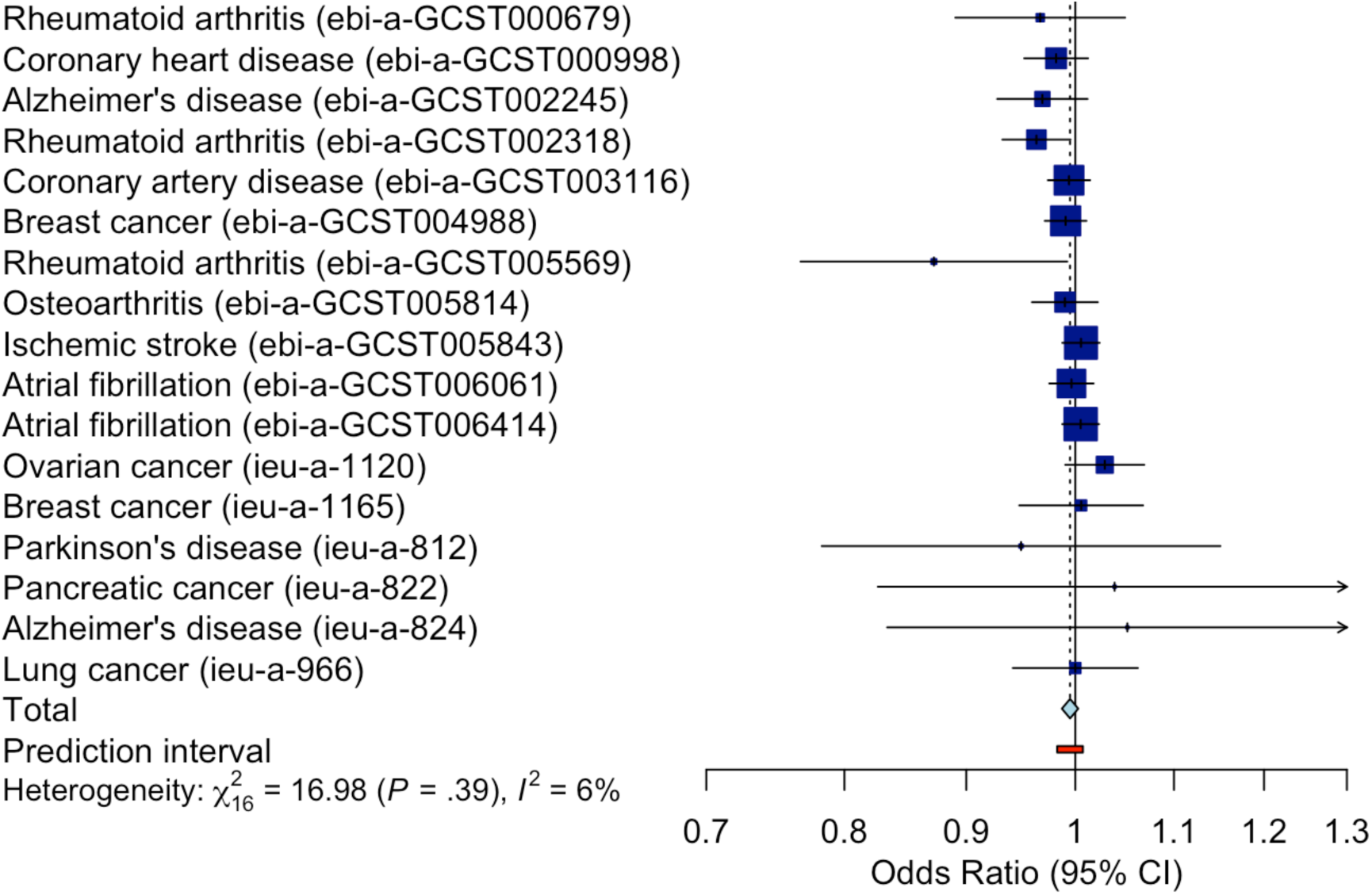
Forest plot of IEAA-associated MR effect sizes (in terms of ORs) for each of the 17 GWAS spanning 11 ARDs (one point for each GWAS) and when metanalysed using a random effect model (in red). The bars represent the corresponding 95% CI.

**Figure S2.**
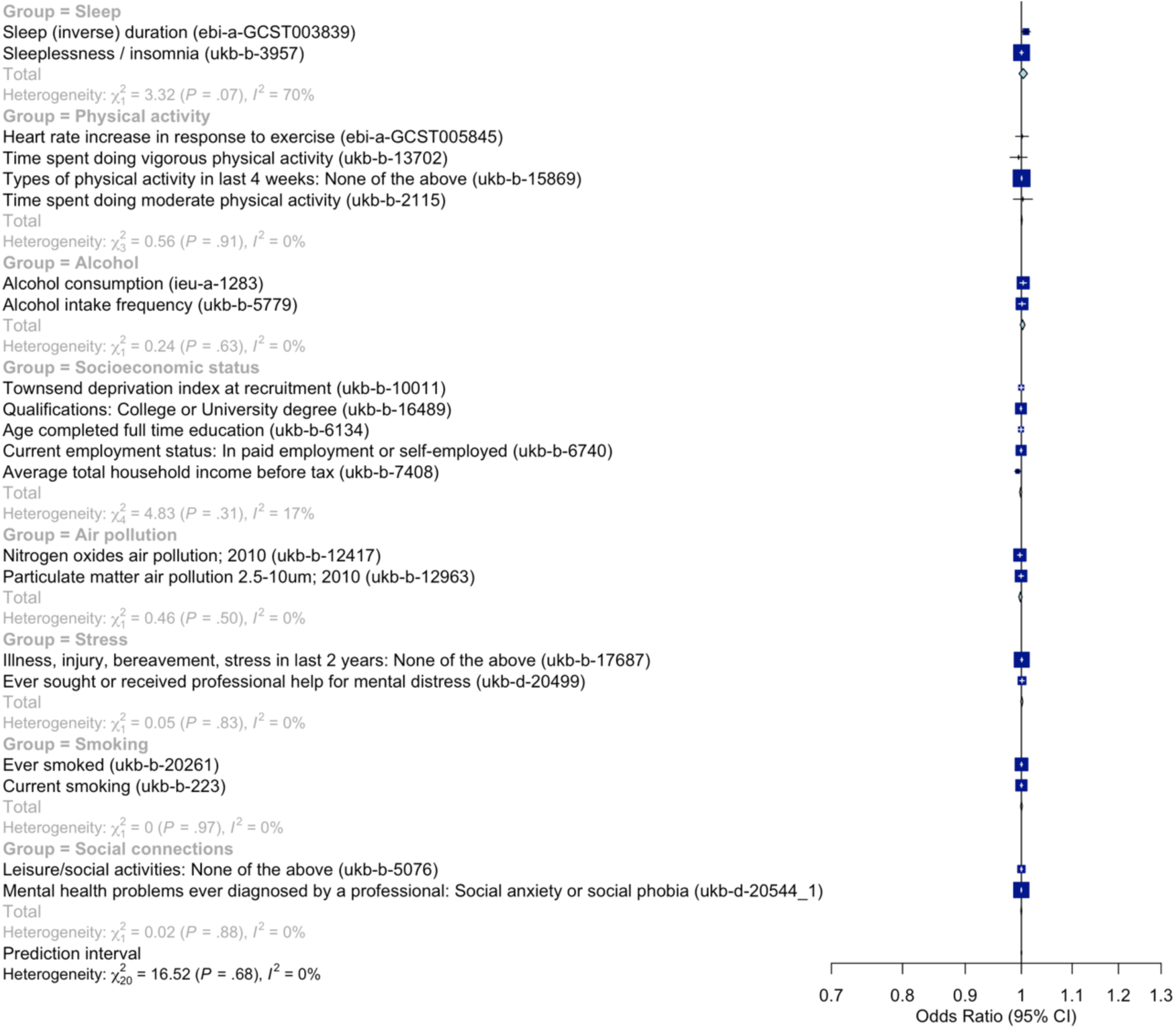
Forest plot of rs2736099-associated OR for environmental, social or lifestyle-related risk factors of ARDs analyzed individually or metanalysed per risk factor group using a random effect model. The bars represent the corresponding 95% CI.

**Figure S3.**
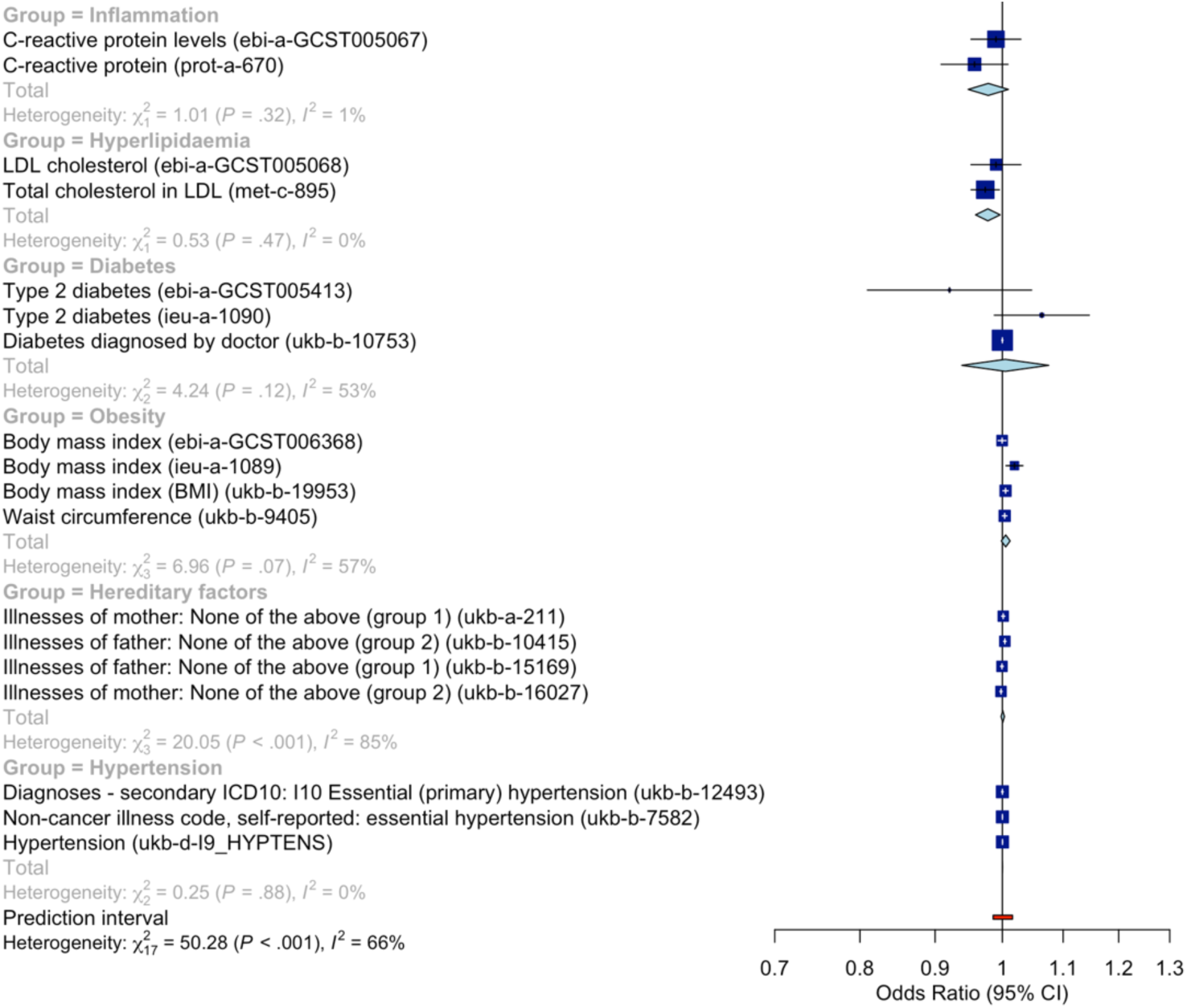
Forest plot of rs2736099-associated OR for hereditary factors and physiological/physical risk factors analyzed individually or metanalysed per risk factor group using a random effect model. Note “group 1 “ refers to heart disease, stroke, high blood pressure, chronic bronchitis/emphysema, Alzheimer’s disease/dementia, and diabetes while “group 2” refers to Parkinson’s disease, severe depression, lung cancer, bowel cancer, and prostate cancer. The bars represent the corresponding 95% CI.

**Table S1.**
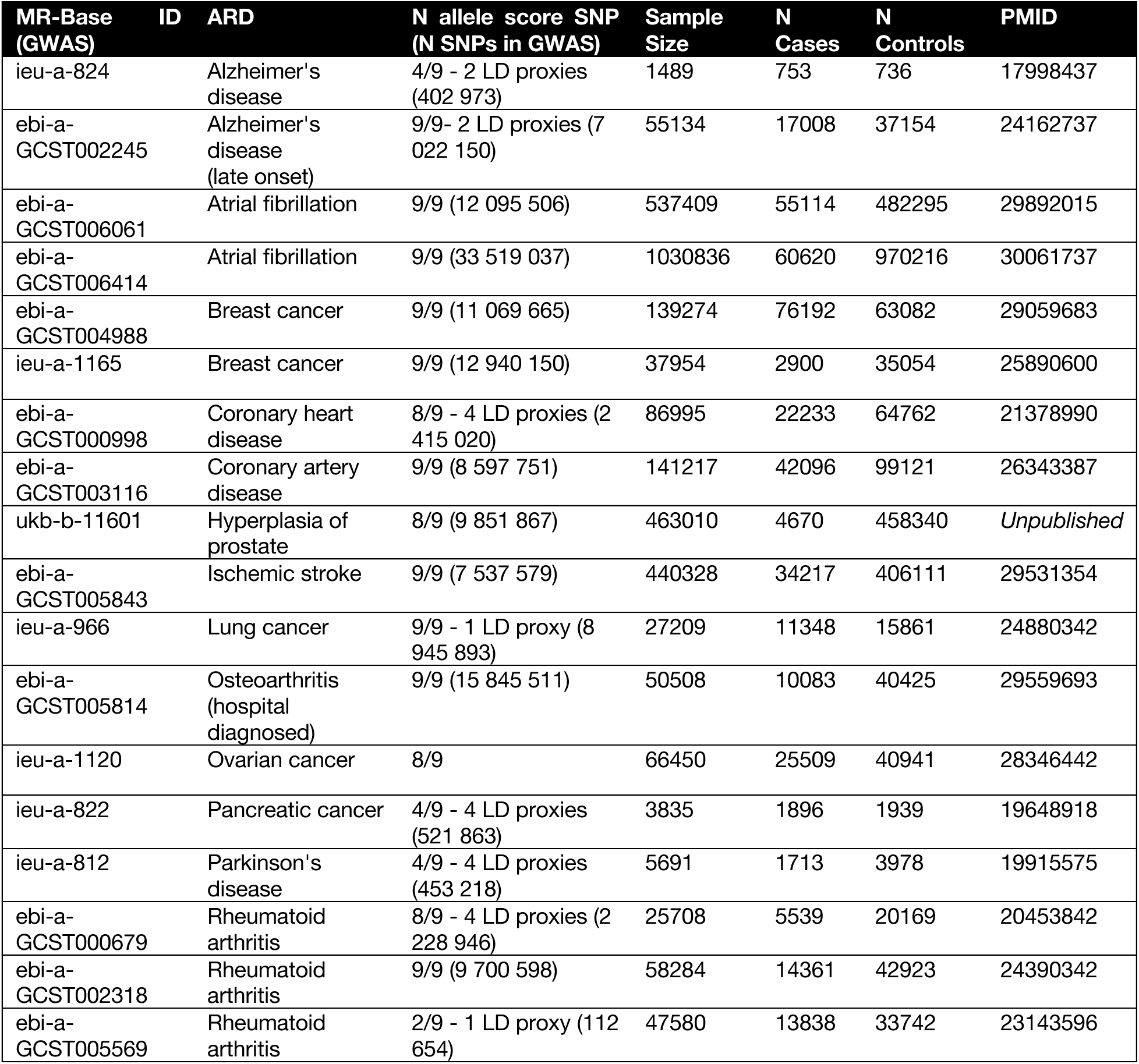
Summary of GWAS studies on ARDs included in the study.

**Table S2.** Odds-ratio (OR) estimates and 95% confidence interval (CI) for each exposure-outcome pair (e.g. IEAA vs. individual ARD) in a two-sample MR tests using RAPS. *On 17 GWAS (Study ukb-b-11601 on benign prostatic hyperplasia was excluded due to its excessive weight in the metanalysis model.)

| ARD | N SNPs | OR | 95% CI | p |
| --- | --- | --- | --- | --- |
| Alzheimer's disease (ebi-a-GCST002245) | 6 | 0.969 | (0.927-1.012) | 0.153 |
| Alzheimer's disease (ieu-a-824) | 2 | 1.052 | (0.834-1.326) | 0.671 |
| Atrial fibrillation (ebi-a-GCST006061) | 6 | 0.996 | (0.975-1.018) | 0.731 |
| Atrial fibrillation (ebi-a-GCST006414) | 8 | 1.005 | (0.987-1.023) | 0.570 |
| Benign prostatic hyperplasia (ukb-b-11601) | 7 | 1.000 | (0.999-1.001) | 0.708 |
| Breast cancer (ebi-a-GCST004988) | 8 | 0.991 | (0.971-1.011) | 0.360 |
| Breast cancer (ieu-a-1165) | 8 | 1.006 | (0.947-1.068) | 0.853 |
| Coronary heart disease (ebi-a-GCST000998) | 7 | 0.982 | (0.952-1.012) | 0.237 |
| Coronary artery disease (ebi-a-GCST003116) | 8 | 0.994 | (0.974-1.014) | 0.560 |
| Ischemic stroke (ebi-a-GCST005843) | 8 | 1.005 | (0.988-1.024) | 0.553 |
| Lung cancer (ieu-a-966) | 8 | 1.000 | (0.941-1.062) | 0.999 |
| Osteoarthritis (ebi-a-GCST005814) | 8 | 0.990 | (0.959-1.022) | 0.534 |
| Ovarian cancer (ieu-a-1120) | 7 | 1.029 | (0.99-1.069) | 0.144 |
| Pancreatic cancer (ieu-a-822) | 2 | 1.039 | (0.826-1.306) | 0.745 |
| Parkinson's disease (ieu-a-812) | 2 | 0.949 | (0.783-1.15) | 0.593 |
| Rheumatoid arthritis (ebi-a-GCST000679) | 5 | 0.967 | (0.891-1.049) | 0.418 |
| Rheumatoid arthritis (ebi-a-GCST002318) | 6 | 0.963 | (0.932-0.995) | 0.025 |
| Rheumatoid arthritis (ebi-a-GCST005569) | 1 | 0.872 | (0.767-0.992) | 0.038 |
| Metanalysis* |  | 0.995 | (0.987-1.003) | 0.207 |

**Table S3.** Odds-ratio (OR) estimates and 95% confidence interval (CI) for the association between rs2736099 and 14 risk factors of ARDs as metanalysed across 39 GWAS. Heterogeneity metrics are also reported in terms of τ^2^, τ, Q, and *I*^2^ statistics. Key: *k* - number of GWAS.

| Risk factor | k | OR | 95%-CI | $\tau^2$ | $\tau$ | Q | $I^2$ | p |
| --- | --- | --- | --- | --- | --- | --- | --- | --- |
| Sleep | 2 | 1.0036 | [0.9957; 1.0115] | <0.0001 | 0.0049 | 3.32 | 69.90% | 0.3766 |
| Hyperlipidaemia | 2 | 0.9776 | [0.9588; 0.9967] | 0 | 0 | 0.53 | 0.00% | 0.0217 |
| Stress | 2 | 1.0008 | [0.9989; 1.0027] | 0 | 0 | 0.05 | 0.00% | 0.3970 |
| Hypertension | 3 | 1.0002 | [1.0000; 1.0004] | 0 | 0 | 0.25 | 0.00% | 0.1259 |
| Inflammation | 2 | 0.9782 | [0.9478; 1.0096] | <0.0001 | 0.0023 | 1.01 | 0.90% | 0.1710 |
| Obesity | 4 | 1.0053 | [0.9984; 1.0123] | <0.0001 | 0.0062 | 6.96 | 56.90% | 0.1351 |
| Socioeconomic status | 5 | 0.9988 | [0.9968; 1.0007] | <0.0001 | 0.0015 | 4.83 | 17.10% | 0.2175 |
| Air pollution | 2 | 0.9982 | [0.9951; 1.0014] | 0 | 0 | 0.46 | 0.00% | 0.2686 |
| Alcohol | 2 | 1.0023 | [0.9980; 1.0068] | 0 | 0 | 0.24 | 0.00% | 0.2953 |
| Physical activity | 4 | 1.0005 | [0.9994; 1.0015] | 0 | 0 | 0.56 | 0.00% | 0.3849 |
| Hereditary | 4 | 1.0007 | [0.9981; 1.0033] | <0.0001 | 0.0024 | 20.05 | 85.00% | 0.6212 |
| Social connections | 2 | 0.9999 | [0.9990; 1.0007] | 0 | 0 | 0.02 | 0.00% | 0.7865 |
| Smoking | 2 | 1.0002 | [0.9986; 1.0017] | 0 | 0 | 0 | 0.00% | 0.8389 |
| Diabetes | 3 | 1.0046 | [0.9384; 1.0755] | 0.0024 | 0.049 | 4.24 | 52.80% | 0.8953 |

**Table S4.** SNPs associated with IEAA in Gibson et al. (2019) (N = 13 493 subjects). Key: SE - Standard error; EAF - Effect allele frequency.

| Phenotype | SNP | Beta | SE | Effect allele | Other allele | EAF | p | Gene |
| --- | --- | --- | --- | --- | --- | --- | --- | --- |
| IEAA (Horvath's clock) | rs76244256 | -1.341 | 0.127 | T | C | 0.046 | 6.23E-26 | TPMT |
| IEAA (Horvath's clock) | rs62078811 | -0.369 | 0.065 | A | G | 0.218 | 1.16E-08 | STXBP4 |
| IEAA (Horvath's clock) | rs388649 | -0.338 | 0.055 | A | T | 0.495 | 6.05E-10 | PIK3CB |
| IEAA (Horvath's clock) | rs1011267 | -0.327 | 0.054 | A | G | 0.503 | 1.58E-09 | C1orf112 |
| IEAA (Horvath's clock) | rs10778517 | 0.335 | 0.054 | T | G | 0.565 | 4.46E-10 | RP11-412D9.4/TMEM263 |
| IEAA (Horvath's clock) | rs4712953 | 0.346 | 0.059 | A | T | 0.725 | 3.60E-09 | SCGN |
| IEAA (Horvath's clock) | rs2736099 | 0.373 | 0.061 | A | G | 0.367 | 8.58E-10 | TERT |
| IEAA (Horvath's clock) | rs7744541 | 0.439 | 0.055 | A | T | 0.418 | 1.93E-15 | NHLRC1 |
| IEAA (Horvath's clock) | rs6440667 | 0.44 | 0.075 | C | G | 0.161 | 4.28E-09 | LINC01214 |
| IEAA (Horvath's clock) | rs79070372 | 0.505 | 0.087 | A | G | 0.111 | 6.07E-09 | GATA2-AS1 |

**Table S5.** GWAS studies in MR-Base in which ARD were investigated as traits.

| MR-Base ID | ARD | Non-independent populations |
| --- | --- | --- |
| ebi-a-GCST001592 | Osteoarthritis | TRUE |
| ebi-a-GCST005811 | Osteoarthritis | TRUE |
| ebi-a-GCST005814 | Osteoarthritis | FALSE |
| ieu-a-1170 | Osteoarthritis | TRUE |
| ebi-a-GCST000679 | Rheumatoid arthritis | FALSE |
| ebi-a-GCST002318 | Rheumatoid arthritis | FALSE |
| ebi-a-GCST005569 | Rheumatoid arthritis | FALSE |
| ieu-a-833 | Rheumatoid arthritis | TRUE |
| ieu-a-834 | Rheumatoid arthritis | TRUE |
| ieu-a-831 | Rheumatoid arthritis | TRUE |
| ebi-a-GCST006061 | Atrial fibrillation | FALSE |
| ebi-a-GCST006414 | Atrial fibrillation | FALSE |
| ebi-a-GCST004296 | Atrial fibrillation | FALSE |
| ieu-a-6 | Coronary heart disease | FALSE |
| ieu-a-7 | Coronary heart disease | FALSE |
| ieu-a-8 | Coronary heart disease | TRUE |
| ieu-a-9 | Coronary heart disease | FALSE |
| ebi-a-GCST000998 | Coronary heart disease | FALSE |
| ieu-a-1108 | Ischemic stroke | TRUE |
| ukb-b-6358 | Ischemic stroke | TRUE |
| ebi-a-GCST005843 | Ischemic stroke | FALSE |
| ukb-d-20002_1278 | Cataract | TRUE |
| ukb-b-11601 | Benign prostatic hyperplasia | FALSE |
| ieu-a-1126 | Breast cancer | TRUE |
| ieu-a-1165 | Breast cancer | FALSE |
| ukb-d-C3_BREAST_3 | Breast cancer | TRUE |
| ukb-d-C3_COLON | Colon cancer | TRUE |
| ieu-a-966 | Lung cancer | FALSE |
| ukb-d-C3_PRIMARY_LYMPHOID_HEMATOPOIETIC | Lymphoma/Leukemia | TRUE |
| ieu-a-1120 | Ovarian cancer | FALSE |
| ukb-b-2160 | Prostate cancer | TRUE |
| ebi-a-GCST002245 | Alzheimer's disease | FALSE |
| ieu-a-298 | Alzheimer's disease | TRUE |
| ieu-a-824 | Alzheimer's disease | FALSE |
| ieu-a-812 | Parkinson's disease | FALSE |
| ukb-b-12141 | Osteoporosis | TRUE |
| ebi-a-GCST004988 | Breast cancer | FALSE |
| ieu-a-822 | Pancreatic cancer | FALSE |
| ieu-a-1082 | Thyroid cancer | FALSE |
| ieu-a-1057 | Gallbladder cancer | FALSE |
| ieu-a-798 | Myocardial infarction | FALSE |
| ebi-a-GCST005195 | Coronary heart disease | FALSE |
| ebi-a-GCST003116 | Coronary heart disease | FALSE |

**Table S6**. Harmonized exposure-outcome datasets. Key: ARD - Age-related disease; SNP - Single nucleotide polymorphism; EAF - Effect allele frequency; LD - Linkage disequilibrium.

*Provided as separate Microsoft Excel file*.

